# In severe first episode major depressive disorder, psychosomatic, chronic fatigue syndrome, and fibromyalgia symptoms are driven by immune activation and increased immune-associated neurotoxicity

**DOI:** 10.1101/2023.08.06.23293708

**Authors:** Michael Maes, Abbas F Almulla, Bo Zhou, Ali Abbas Abo Algon, Pimpayao Sodsai

## Abstract

**Background:** Major depressive disorder (MDD) is accompanied by activated neuro-immune pathways, increased physiosomatic and chronic fatigue-fibromyalgia (FF) symptoms. The most severe MDD phenotype, namely major dysmood disorder (MDMD), is associated with adverse childhood experiences (ACEs) and negative life events (NLEs) which induce cytokines/chemokines/growth factors.

**Aims:** To delineate the impact of ACE+NLEs on physiosomatic and FF symptoms in first episode (FE)-MDMD, and examine whether these effects are mediated by immune profiles.

**Methods:** ACEs, NLEs, physiosomatic and FF symptoms, and 48 cytokines/chemokines/growth factors were measured in 64 FE-MDMD patients and 32 normal controls.

**Results:** Physiosomatic, FF and gastro-intestinal symptoms belong to the same factor as depression, anxiety, melancholia, and insomnia. The first factor extracted from these seven domains is labeled the physio-affective phenome of depression. A part (59.0%) of the variance in physiosomatic symptoms is explained by the independent effects of interleukin (IL)-16 and IL-8 (positively), CCL3 and IL-1 receptor antagonist (inversely correlated). A part (46.5%) of the variance in physiosomatic (59.0%) symptoms is explained by the independent effects of interleukin (IL)-16, tumor necrosis factor-related apoptosis-inducing ligand (TRAIL) (positively) and combined activities of negative immunoregulatory cytokines (inversely associated). Partial Least Squares analysis shows that ACE+NLEs exert a substantial influence on the physio-affective phenome which are partly mediated by an immune network composed of IL-16, CCL27, TRAIL, macrophage-colony stimulating factor, and stem cell growth factor.

**Conclusions:** The physiosomatic and FF symptoms of FE-MDMD are partly caused by immune-associated neurotoxicity due to T helper (Th)-1 polarization, Th-1, and M1 macrophage activation and relative lowered compensatory immunoregulatory protection.

## Introduction

Using a novel precision psychiatry methodology that incorporates machine learning techniques, we recently demonstrated that there are two qualitatively distinct phenotypes of major depressive disorder (MDD), namely Major Dysmood Disorder (MDMD) and Simple Dysmood Disorder (SDMD) (Maes, Moraes et al. 2021, Maes, Rachayon et al. 2022, Maes and Almulla 2023). MDMD is distinguished from SDMD by a higher recurrence of illness (ROI), which is derived from the number of depressive episodes and the number of lifetime suicidal attempts and ideation (Maes, Moraes et al. 2019). In addition, patients with MDMD score higher on lifetime and current suicidal behaviors and attempts, cognitive deficits, and disabilities, than those with SDMD (Maes, Moraes et al. 2021, Maes and Almulla 2023). In contrast to SDMD, MDMD is associated with elevated levels of pure depressive symptoms (such as depressed mood, feelings of guilt, suicidal ideation, and decreased interest) (Hamilton 1960), pure anxiety symptoms (anxious mood, tension, fears, and anxious behavior observed during the interview) (Hamilton 1959), insomnia (early, middle, and late), symptoms of melancholia (including psychomotor retardation, psychomotor agitation, diurnal variation, and early awakening), and total sums on both the Hamilton Depression (HAMD) and Anxiety (HAMA) Rating Scales (Hamilton 1959, Hamilton 1960, Maes, Moraes et al. 2021, Maes and Almulla 2023).

However, rating scales such as the HAMD and HAMA score high on psychosomatic symptoms, including anxiety somatic, somatic gastro-intestinal, general somatic, genital symptoms, and hypochondriasis of the HAMD (Hamilton 1960), and somatic anxiety, sensory, cardiovascular, gastrointestinal (GIS), genitourinary, and respiratory symptoms of the HAMA (Hamilton 1959). Multiple studies have consistently demonstrated that psychosomatic symptoms play a significant role in both “endogenous” depression, and depression resulting from medical conditions (Anderson, Maes et al. 2012, Maes, Kubera et al. 2013, Al-Hakeim, Hadi et al. 2022, Al-Hakeim, Al-Naqeeb et al. 2023). Nevertheless, due to the discovery that the psychosomatic symptoms of depression have a biological basis (as evidenced below), we have chosen to redefine this collection of symptoms as “physiosomatic” (Anderson et al., 2012). A comparative analysis revealed a notable disparity in the intensity of physiosomatic symptoms between individuals diagnosed with MDMD and the control group (Maes, Moraes et al. 2021, Maes, Rachayon et al. 2022, Maes, Moraes et al. 2023). Additional physiosomatic symptoms that resemble those found in chronic fatigue syndrome (CFS) and fibromyalgia can be evaluated through the utilization of the Fibro-Fatigue (FF) scale (Zachrisson, Regland et al. 2002). These symptoms include muscle pain, muscle tension, fatigue, autonomic and gastrointestinal symptoms, headache, and a flu-like malaise (Maes, Ringel et al. 2013, Morris, Puri et al. 2019). Nevertheless, there is a lack of available data regarding the potential increase in FF symptoms in individuals with MDMD compared to those with SDMD.

Significantly, individuals diagnosed with MDMD and those experiencing their first episode of MDMD (FE-MDMD) exhibit notable elevations in neuro-immune, autoimmune, and gut-brain pathways. These pathways encompass specific modifications in the cytokine network, gut microbiome, gut dysbiosis, heightened translocation of Gram-negative bacteria, increased atherogenicity, and diminished antioxidant, and neurotrophic defenses (Simeonova, Stoyanov et al. 2021, Maes 2022, Maes, Vasupanrajit et al. 2023). The neuro-immune disorders observed in MDD can be attributed solely to the neuro-immune abnormalities present in MDMD, while there is no association between activated immune-inflammatory pathways and SDMD (Maes, Rachayon et al. 2022, Almulla, Ali Abbas Abo et al. 2023a). The immune characteristics that are of significance in MDMD encompass the activation of classical M1 macrophages, alternative M2 macrophages, T helper-1 (Th-1) cells, and Th-17 cells. These immune profiles lead to the activation of the immune-inflammatory response system (IRS) and an increase in the levels of neurotoxic (NT) immune products (Maes, Rachayon et al. 2022, Almulla, Ali Abbas Abo et al. 2023a). According to the study conducted by Almulla et al. (2023), the activation of Th-1 cells and the increased presence of interleukin (IL)-16, tumor necrosis factor (TNF)-α, and TNF-related apoptosis-inducing ligand (TRAIL) are responsible for inducing IRS activation in MDMD. The augmented immune-associated neurotoxicity (Immu-NT) observed in individuals with MDMD is predominantly attributed to heightened concentrations of IL-16, TNF-α, IL-6, CCL2, CCL11, CXCL1, and CXCL10 (Almulla, Ali Abbas Abo et al. 2023a). On the other hand, individuals diagnosed with MDMD demonstrate heightened concentrations of immune substances that possess negative immunoregulatory or anti-inflammatory properties, including IL-10, IL-4, soluble IL-1 receptor antagonist (sIL-1RA), and sIL-2R (Almulla, Ali Abbas Abo et al. 2023a). The relationship between depression, as a construct derived from measures such as HAMD, HAMA, and other depression scores, and activated immune profiles has been found to be highly predictive. However, it remains unclear whether the neuro-immune pathways responsible for MDMD are associated with physiosomatic and FF symptoms in MDMD.

It is widely recognized that experiencing multiple adverse childhood experiences (ACEs) and negative life events (NLEs) is associated with an elevated risk of developing MDD, anxiety disorders, and suicidal behaviors in adulthood (Paykel, Emms et al. 1980, Kendler, Karkowski et al. 1999, Kraaij, Arensman et al. 2002, Agnew-Blais and Danese 2016, Maes, Congio et al. 2018, Maes, Moraes et al. 2019, Abe, Sirichokchatchawan et al. 2022, Maes, Rachayon et al. 2022). Emerging research suggests that there is a significant association between ACEs, the combination of ACEs and NLEs within the past year, and the development of MDD, with a particular emphasis on FE-MDMD (Maes and Almulla 2022). These factors have been found to strongly predict various aspects of MDD, such as the severity of depression and anxiety symptoms, lifetime history and current presence of suicidal behaviors, as well as physiosomatic symptoms (Maes, Rachayon et al. 2022, Almulla, Ali Abbas Abo et al. 2023a). The association between the occurrence of FE-MDMD and current suicidal behaviors is primarily linked to the cumulative impact of ACEs and NLEs (Maes and Almulla 2023).

Furthermore, the occurrence of ACEs is strongly correlated with the activation of immune-inflammatory cytokines, chemokines and growth factor responses, oxidative stress reactions, alterations in the gut microbiome, and diminished antioxidant and neurotrophic responses in later stages of life (Maes, Moraes et al. 2019, Maes, Moraes et al. 2021, Maes, Vasupanrajit et al. 2023, Almulla, Ali Abbas Abo et al. 2023b). In addition, ACEs have been found to be linked with the activation of M1 and T cytokines, polarization towards a Th-1 response, heightened expression of T cell activation markers such as CD4, CD40L, and CD71, as well as increased IL-16 signaling. Moreover, ACEs have been associated with elevated levels of chemokines including CCL2, CCL5, CCL27, CXCL9, and CXCL10, as well as growth factors such as stem cell factor (SCF), stem cell growth factor (SCGF), hepatic growth factor (HGF), and platelet-derived growth factor (PDGF) (Maes, Vasupanrajit et al. 2023, Almulla, Ali Abbas Abo et al. 2023b). Furthermore, the effects of ACE+NLEs on the depression phenotype are mediated by activated neuroimmune pathways and Th-1 cell activation (Maes, Vasupanrajit et al. 2023).

There is a substantial body of literature indicating that the physiosomatic and FF symptoms associated with MDD may be explained by the activation of neuro-immune pathways in both the peripheral and central nervous systems. The pathways involved in this context encompass M1 and Th-1 pathways, along with the participation of distinct immune molecules, namely IL-6, IL-1β, and TNF-α (Maes 2011, Maes, Mihaylova et al. 2012, Morris and Maes 2012, Anderson, Berk et al. 2014, Maes, Anderson et al. 2014, Morris, Berk et al. 2014).

The specific mechanisms through which ACEs or ACE+NLEs could impact the physiosomatic and FF symptoms of MDD/MDMD remain uncertain. It is unclear whether these effects are mediated by the activation of the IRS, specific immune phenotypes like Th-1 or M1 phenotypes, or specific cytokines, chemokines, and growth factors such as IL-6, IL-1β, and TNF-α. Therefore, the precise hypotheses of this study are as follows: the impact of ACEs and NLEs on physiosomatic and FF symptoms is influenced by increased activity of the IRS and Immu-NT, specifically through the release of certain M1 cytokines such as IL-6, IL-1β, and TNF-α.

## Materials and methods

### Participants

In accordance with the Diagnostic and Statistical Manual of Mental Disorders, Fifth Edition (DSM-5) (American Psychiatric Association 2013), a senior psychiatrist at Alhakiem Hospital in Najaf, Iraq, identified and recruited 64 individuals with FE-MDD and 32 healthy controls. All patients diagnosed with MDD were observed to be in the acute phase of the disease, with no indications of complete or partial remission. It should be noted that all FE-MDD patients met the diagnostic criteria for MDMD as detailed by Maes, Vasupanrajit, et al. (2023) (Maes, Vasupanrajit et al. 2023). In addition, the senior psychiatrist recruited healthy controls from the same geographical area to serve as the control group. This group included hospital staff and patients’ acquaintances. All participants were enlisted between October 2021 and March 2022.

Exclusions were made for those with chronic liver or kidney disease, women who were pregnant or nursing, and those with a history of multiple sclerosis, Parkinson’s disease, stroke, or Alzheimer’s disease. Similarly, psoriasis, rheumatoid arthritis, inflammatory bowel disease, cancer, type 1 diabetes, and scleroderma exhibited the same pattern. In addition, we excluded participants who had experienced an acute COVID-19 infection, severe or critical COVID-19 disease, Long COVID, or a COVID-19 infection within the previous six months. Our study did not include individuals taking immunosuppressive or immunomodulatory medications, nor did it include those taking therapeutic doses of antioxidants or omega-3 supplements (last three months).

The process of meticulously selecting the control group was accorded significant consideration. Excluded from the control group were participants with a documented lifetime history of clinical depression or dysthymia, a family history of depression, mania, psychosis, or substance use disorder, or a history of suicide. Dysthymia (excluding cases of double depression), schizophrenia, schizoaffective disorder, bipolar disorder, autism spectrum disorders, substance use disorders (excluding nicotine dependence), post-traumatic stress disorder, psycho-organic disorders, generalized anxiety disorder, and obsessive-compulsive disorder were excluded from the study.

Before participating in the study, all participants, or, if applicable, their parents or legal custodians, provided informed consent in writing. Document No. 18/2021 indicates that the Ethics Committee of the College of Medical Technology at the Islamic University of Najaf in Iraq has approved this investigation. The research was conducted in accordance with both Iraqi and international ethical and privacy regulations. In prominent documents such as the Declaration of Helsinki by the World Medical Association, the Belmont Report, the CIOMS Guideline, and the International Conference on Harmonization of Good Clinical Practice, a variety of non-exhaustive principles are outlined. Our organization’s institutional review board (IRB) is committed to upholding the highest standards of quality, ensuring precise compliance with the International Guideline for the Conduct of Safe Human Research (ICH-GCP).

### Clinical assessments

Using the HAMD (Hamilton 1960), the HAMA (Hamilton 1959), assessments of suicidal behaviors, and evaluations of disease recurrence scores (Maes and Almulla 2023), a diagnosis of MDMD can be established using machine learning techniques. Alternatively, when both the HAMD and the HAMA exceed twenty-two points, it may also be diagnosed. As such, the current study included only patients with FE-MDMD. The physical, mental, and behavioral health of the participants was evaluated by a senior psychiatrist who used a systematic interview and standardized procedures. The senior psychiatrist collected sociodemographic, clinical, and psychological information through semi-structured interviews.

The same professional utilized the HAMD and the HAMA to evaluate the severity of depression and anxiety, respectively. In the current study, all physiosomatic symptoms of the HAMD and HAMA were excluded to compute pure depression and pure anxiety scores, respectively. The former concept was conceptualized as a collection of symptoms including depressed mood, feelings of guilt, suicidal ideation, and decreased interest. The score for pure anxiety was determined by adding the scores for anxious mood, tension, fears, and anxious behavior observed during the interview. The physiosomatic symptom score was calculated as a z unit-based composite score based on the sum of the z scores of HAMD and HAMA physiosomatic symptoms, namely anxiety somatic, somatic gastrointestinal, and genitourinary symptoms, hypochondriasis, somatic sensory, cardiovascular, gastrointestinal, genitourinary, autonomic, and respiratory symptoms. The pure FF symptoms were calculated as the sum of the FF items after exclusion of non-physiosomatic symptoms: muscle pain, muscle tension, fatigue, autonomic, gastro-intestinal symptoms, headache, and a flu-like malaise (all FF scale items after exclusion of all non-physiosomatic symptoms, including cognitive deficits, and sadness). GIS symptoms were conceptualized as a z unit-based composite score based on symptoms of the HAMA and FF scores, namely somatic gastrointestinal and GIS symptoms. Insomnia was conceptualized as a z value-based composite, namely sum of insomnia (HAMA item), insomnia early + insomnia middle + insomnia late (HAMD items) + sleep disorders (FF). Melancholia was conceptualized as a composite based on the z scores of insomnia late, psychomotor retardation, psychomotor agitation, loss of weight, and diurnal variation. This study utilized two items of the Columbia Suicide Severity Rating Scale (C-SSRS) to evaluate suicidal behaviors: the quantification of suicide attempts within the previous year and the assessment of the frequency of suicidal ideation within the previous three months (Posner, Brown et al. 2011). A composite score of suicidal behaviors (SB) was calculated by adding the weighted z scores for the HAMD suicide item, the number of suicide attempts, and the frequency of suicidal ideation.

The ACEs Questionnaire (Felitti, Anda et al. 1998) was used to measure the extent of adverse childhood experiences. There are a total of twenty-eight items on the scale, which comprise the scoring of ten distinct domains. These include (1) mental trauma, (2) physical trauma, (3) sexual abuse, (4) mental neglect, (5) physical neglect, (6) witnessing domestic violence involving the mother, (7) presence of a family member with drug abuse issues, (8) presence of a family member with depression or mental illness, (9) experiencing the loss of a parent due to separation, death, or divorce, and (10) having an incarcerated family member.

Various ACE scores were calculated, including the total sum of all ACEs (termed total ACE), the sum of physical trauma, mental neglect, and family member with substance abuse (termed ACE247), and the sum of ACE247 and a family member in prison (termed ACE24710). In addition, the NLEs scale was used to assess the incidence of NLEs in the preceding year (Buri 2018). For the purposes of this investigation, the following factors were considered: a serious accident, the death of a family member or close friend, divorce or separation, unemployment, job loss, alcohol-related issues, drug-related issues, witnessing physical altercations or assaults, experiences of abuse or violent crime, encounters with law enforcement difficulties, problem gambling, familial incarceration, overcrowding in the household, and instances of discrimination. Consequently, we performed calculations to ascertain the combined effects of ACE and the occurrence of one or more NLEs, which we referred to as ACE+NLEs. Body mass index (BMI) was calculated by dividing the participants’ weight in kilograms by the square of their height in meters. Using the diagnostic criteria specified in the fifth edition of the Diagnostic and Statistical Manual of Mental Disorders (DSM-5), tobacco use disorder was evaluated.

### Biochemical assays

In this investigation, a disposable syringe and serum tubes were used to obtain a 5 mL venous blood sample from each participant while they were fasting. Blood samples were collected between 8:00 and 11:00 in the morning. The serum and blood cells were effectively separated following centrifugation at 35,000 revolutions per minute (rpm). The serum was then transferred into small Eppendorf containers and stored at -80°C until its use in the biomarker assays was deemed necessary. Bio-Plex Multiplex Immunoassay kits (Bio-Rad Laboratories Inc., Hercules, USA) were used to measure the levels of forty-eight cytokines/chemokines/growth factors in the serum of all participants. For measuring the concentrations of these proteins in blood serum, we employed a fluorescence-based method. The researchers measured the fluorescence intensity (FI) of each protein and used the values obtained after subtracting the blank value (Rachayon, Jirakran et al. 2022, Almulla, Ali Abbas Abo et al. 2023a). The Electronic Supplementary File (ESF) comprises Table 1, which provides an exhaustive listing of the analytes identified by our research. In addition, each analyte’s respective gene ID and alternate names are listed in this table. The coefficients of variation (CV) for all analytes in the assay were less than 11.0%. To determine the concentrations of the analytes, we employed the manufacturer-supplied standards. Subsequently, the out-of-range (OOR) concentration rate was calculated, i.e., the proportion of concentrations that fell below the minimum detectable level. In over 80% of cases, cytokines/chemokines/growth factors with concentrations below the lowest out-of-range (OOR) value were excluded from the statistical analysis. The aforementioned cytokines, including IFN-α2, IL-3, IL-7, and IL-12p40, were therefore excluded from our study. As dummy variables, we accounted for analytes with quantifiable prevalence rates between 20% and 40% when calculating our composite scores. As a result, diverse immune profiles were computed, as described in Table 2 of the ESF. These profiles covered all analytes with the exception of those with detectable concentrations below 20% (Rachayon, Jirakran et al. 2022, Thisayakorn, Thipakorn et al. 2022, Almulla, Ali Abbas Abo et al. 2023a). Table 2 of the ESF lists the variables used to generate M1, M2, M1/M2 (z M1 - z M2), Th1, Th2, Th1/Th2 (z Th-1 - z Th-2), Th-17, IRS, CIRS, IRS/CIRS (z IRS - z CIRS), and Immu-NT profiles. In addition, the individual cytokines, chemokines, and growth factors were analyzed to ascertain whether there were significant differences in immune profiles between the study groups.

**Table 1.** Sociodemographic and clinical data in healthy controls and patients with major dysmood disorder (MDMD) with very severe physiosomatic symptoms and immune profiles (Severe MDMD) and MDMD patients with milder physiosomatic and immune scores (Milder MDMD).

| Variables | HC a<br>n=32 | Milder MDMD b<br>n=29 | Severe MDMD c<br>n=35 | F/X<sup>2</sup> | df | p |
| --- | --- | --- | --- | --- | --- | --- |
| Age (years) | 32.0 ±7.6 | 33.5 ±9.2 | 29.8 ±8.3 | 1.64 | 2/93 | 0.200 |
| Sex (M/F) | 23/9 | 18/11 | 23/12 | 0.68 | 2 | 0.711 |
| Educated (No/Yes) | 11/21 | 9/20 | 10/25 | 0.26 | 2 | 0.877 |
| Single/married | 15/17 | 17/12 | 22/13 | 1.83 | 2 | 0.401 |
| BMI (kg/m2) | 26.19 ±4.11 | 24.74 ±3.45 | 34.24 ±3.77 | 2.33 | 2/93 | 0.103 |
| Smoking (No/Yes) | 14/18 | 22/7 | 28/7 | 11.47 | 2 | 0.003 |
| Previous COVID (No/yes) | 20/12 | 18/11 | 22/13 | 0.00 | 2 | 0.998 |
| Duration of illness (months) | - | 2.26 ±2.78 | 2.58 ±2.84 | 0.21 | 1/62 | 0.649 |
| Pure depression | 0.47 ±0.76 b,c | 10.62 ±1.54 a | 10.82 ±1.48 a | 654.52 | 2/93 | <0.001 |
| Pure anxiety | 0.87 ±0.87 b,c | 9.17 ±1.28 a | 9.20 ±1.47 a | 478.54 | 2/93 | <0.001 |
| Pure FF | 1.16 ±0.92 b,c | 8.90 ±4.03 a | 9.31 ±3.06 a | 79.06 | 2/93 | <0.001 |
| Physiosomatic symptoms | -1.275 ±0.325 b,c | 0.374 ±0.633 a,c | 0.673 ±0.558 a,b | 132.46 | 2/93 | <0.001 |
| Melancholia | -1.220 ±0.358 b,c | 0.238 ±0.541 a,c | 0.658 ±0.690 a,b | 104.21 | 2/93 | <0.001 |
| Insomnia | -1.225 ±0.299 b,c | 0.465 ±0.548 a | 0.613 ±0.752 a | 106.30 | 2/93 | <0.001 |
| GIS symptoms | -1.030 ±0.294 b,c | 0.276 ±0.644 a,c | 0.746 ±0.762 a,b | 76.04 | 2/93 | <0.001 |
| Autonomic symptoms | -0.300 ±0.523 c | -0.136 ±0.889 | 0.280 ±1.302 a | 3.18 | 2/93 | <0.001 |
| Total ACEs | 0.41 ±0.499 b,c | 1.52 ±0.986 a,c | 2.00 ±1.138 a,b | 25.93 | 2/93 | <0.001 |
| ACE+NLEs | -0.994 ±0.552 b,c | 0.343 ±10.705 a | 0.607 ±0.954 a | 41.00 | 2/93 | <0.001 |
| Fluoxetine (No/yes) | - | 18/11 | 23/12 | 0.09 | 1 | 0.762 |
| Amitryptiline (No/yes) | - | 27/2 | 30/5 | FEPT | - | 0.442 |
| Escitalopram (No/yes) | - | 27/2 | 31/4 | FEPT | - | 0.681 |
| Olanzapine (No/yes) | - | 27/2 | 32/3 | FEPT | - | 1.0 |
| Mirtazapine (No/yes) | - | 23/6 | 33/2 | FEPT | - | 0.127 |
| Drug naïve (No/yes) | - | 19/10 | 19/16 | 0.83 | 2 | 0.362 |
| PC_immune+phenome | -1.347 ±0.301 b,c | 0.437 ±0.129 a,c | 0.869 ±0.218 a,b | 849.29 | 2/93 | <0.001 |

All results are shown as mean ±SD. F: results of analysis of variance, $\chi^2$ : results of analysis of contingency analysis; FEPT: Fisher's exact probability test, a,b,c: pairwise comparisons among group means at p=0.05.
FF: Fibro-Fatigue, GIS: gastro-intestinal symptoms, ACE: adverse childhood experiences, NLE: negative life events, PC\_immune+phenome: a principal component extracted from 9 immune variables and 6 phenome symptom domains.

**Table 2.** Immune profiles in healthy controls (HC) and patients with major dysmood disorder (MDMD) with very severe physiosomatic symptoms and immune profiles (Severe MDMD) and MDMD patients with milder physiosomatic and immune scores (Milder MDMD).

| Variables (z scores) | HC<br>n=32 | Milder MDMD<br>n=29 | Severe MDMD<br>n=35 | F/X<sup>2</sup> | df | P |
| --- | --- | --- | --- | --- | --- | --- |
| Classical macrophage M1 | -0.604 ±0.918 b,c | 0.005 ±0.902 a,c | 0.548 ±0.839 a,b | 14.16 | 2/93 | <0.001 |
| Alternative macrophage M2 | -0.479 ±0.851 b,c | 0.043 ±0.914 a | 0.402 ±1.030 a | 7.42 | 2/93 | 0.001 |
| zM1 – zM2 | 0.124 ±0.724 | -0.039 ±0.489 | 0.146 ±0.581 | 1.73 | 2/93 | 0.184 |
| Thelper (Th)-1 | -0.592 ±1.086 b,c | 0.017 ±0.695 a,c | 0.527 ±0.841 a,b | 13.17 | 2/93 | <0.001 |
| Th-2 | -0.398 ±0.975 c | -0.150 ±0.844 c | 0.489 ±0.961 a,b | 8.12 | 2/93 | <0.001 |
| zTh-1 – zTh-2 | -0.194 ±0.605 b | 0.168 ±0.484 a | 0.0380 ±0.454 | 3.86 | 2/93 | 0.025 |
| Th-17 | -0.392 ±1.305 c | -0.098 ±0.700 c | 0.440 ±0.696 a,b | 6.71 | 2/93 | 0.002 |
| IRS | -0.632 ±1.051 b,c | 0.002 ±0.679 a,c | 0.577 ±0.829 a,b | 16.10 | 2/93 | <0.001 |
| CIRS | -0.564 ±0.933 b,c | -0.055 ±0.629 a,c | 0.561 ±1.023 a,b | 13.43 | 2/93 | <0.001 |
| zIRS - zCIRS | -0.163 ±1.002 | 0.136 ±0.977 | 0.037 ±1.025 | 0.71 | 2/93 | 0.494 |
| Immu-NT | -0.575 ±1.108 b,c | -0.043 ±0.665 a,c | 0.560 ±0.820 a,b | 13.71 | 2/93 | <0.001 |

All results are shown as mean ±SD. F: results of analysis of variance.
IRS: immune-inflammatory response system, CIRS: compensatory immunoregulatory system. Immu-NT: immune-associated neurotoxicity.

### Statistics

This study’s statistical analyses were performed with IBM SPSS 29, Windows version. To compare continuous variables between study groups, statistical tests such as analysis of variance (ANOVA) and the Kruskal-Wallis test were utilized. In contrast, nominal variable comparisons were performed using contingency table analysis, specifically the Chi-square test. In addition, Pearson’s and point-biserial correlation coefficients were used to analyze the relationships between scale variables and binary variables. Due to the observed interconnections between cytokines, chemokines, and growth factors within the immune (cytokines and chemokines) and growth factor networks, it was decided not to implement false discovery rate (FDR) p-value correction (Maes, Plaimas et al. 2021, Rachayon, Jirakran et al. 2022). The researchers used manual multiple regression analysis to investigate the impact of ACEs, NLEs, and additional demographic factors on the immune profiles. In a similar manner, these analyses examined the influence of different predictor variables, namely immune profiles, ACEs, and NLEs, on the manifestations of depression. In addition, automatic forward stepwise regressions were employed. A significance level of p=0.05 was used to determine the inclusion of variables, and a significance level of p=0.06 was used to determine their exclusion. This method made it easier to determine which variables should be included in the final regression model and which ones should be excluded. For each variable included in the final regression models, the standardized coefficients, t-statistics, and exact p-values were calculated. In addition, we determined the F statistics, their respective p-values, and the effect magnitude using the partial Eta squared. Using appropriate statistical measures, the presence of multicollinearity and collinearity in the data was extensively examined. A tolerance limit of 0.25 and a threshold for the variance inflation factor of four were used. In addition, for the purpose of this analysis, we utilized the condition index and variance proportions derived from the collinearity diagnostics table. Using the White test and a modified variation of the Breusch-Pagan test, heteroskedasticity was identified. All the preceding analyses employed two-tailed tests. A significance level of 0.05 or lower was regarded as statistically significant. As required, we utilized transformations such as logarithmic, square-root, rank-based inversed normal (RINT), and a Winsorization technique to obtain a normal distribution for our data indicators.

Partial least squares (PLS) analysis was used to investigate the causal relationships between ACEs and NLEs, immunological profiles, and the physiosomatic symptoms of depression. The output variable was a latent vector derived from the diverse symptom domains, and ACE+NLEs and immune profiles as explanatory variables. In addition, the immune profiles were allowed to mediate the effects of ACE+NLEs on the clinical assessment scores. Only when both the external and internal models met the predetermined quality criteria was a comprehensive partial least squares (PLS) analysis conducted. These quality criteria are: a) Confirmatory tetrad analysis (CTA) verifies that the latent vectors derived from the indicators have not been incorrectly specified as reflective models. b) The blindfolding procedure reveals that the cross-validated redundancy of the construct is adequate. c) The latent vectors exhibit strong construct and convergence validity, as indicated by composite reliability values greater than 0.80, Cronbach’s alpha values greater than 0.70, and average variance extracted (AVE) values greater than 0.5. d) At a significance level of p 0.001, all loadings on the extracted latent vectors exceed 0.65. e) The model fit is regarded satisfactory if the standardized root squared residual (SRMR) is less than 0.08. Consequently, a thorough pathway analysis is conducted using PLS-Structural Equation Modeling (PLS-SEM). In the analysis, SmartPLS software and 5,000 bootstrap samples were utilized. Path coefficients and their respective p-values were calculated. In addition, specific indirect effects, total indirect effects (mediated effects), and total effects were calculated if the model quality data met the specified conditions. The estimated minimum sample size is 103 based on a power analysis (conducted with G*Power 3.1.9.4) using a linear multiple regression analysis with an estimated effect size of 0.111 (corresponding to approximately 10% of the variance explained), a significance level (alpha) of 0.05, and statistical power of 0.8, with three covariates.

## Results

### Socio-demographic and clinical data

**Table 1** shows the sociodemographic and clinical data of the participants divided into three groups using a PC extracted from 6 symptom domains (pure_depression, pure_anxiety, physiosomatic, insomnia, melancholia, and gastro-intestinal symptoms), and the major immune profiles (M1, Th-1, IRS, CIRS) and cytokines/chemokines (IL-16, TRAIL, CLC27, SCGF, M-CSF) (labeled: PC_immune+phenome) that differentiate controls from patients. We were able to extract one validated PC from these 15 variables (KMO=0.839, Bartlett’s test of sphericity = 1155.581, df=36, p<0.001, EV=61.5, all loading of the 15 variables are >0.694). Consequently, using a visual binning method the PC score was divided in three groups using -0.606 and +0.608. Table 1 shows the mean values of the PC_immune+phenome scores. There was a strong association between these groups and the clinical diagnosis (controls versus MDMD patients): the PC-derived class with a PC score < -0.606 were the 32 controls, while the patients were divided into those with severe symptoms and immune aberrations (labeled “severe MDMD”) versus milder symptoms and immune disorders (“milder MDMD”).

Table 1 shows that there were no significant differences in age, sex, education, marital status, BMI, and previous COVID-19 infection between the three study groups. Pure_depression, pure_anxiety, pure_FF, and insomnia scores were higher in patients than controls, while physiosomatic, melancholia, and GIS scores were different between the three groups and increased from controls to milder MDMD to severe MDMD. Autonomic symptoms were significantly increased in the latter group as compared to controls. There were no significant differences in the use of amitriptyline, escitalopram, olanzapine, and mirtazapine between the two depression study groups. There were twenty-six drug-naïve patients and the medicated / unmedicated ratio was not significantly different between both depression subgroups.

### Immune profiles and cytokine/chemokine/growth factor measurements

**Table 2** shows the measurements of the immune profiles in controls versus both depression subgroups. Classical M1, Th-1, Th-2, IRS, CIRS and the Immu-NT profiles differed significantly between the three subgroups and increased from controls to the milder MDMD group to severe MDMD. There were no differences in the zM1 – zM2 and zIRS – zCIRS scores between the three groups, whereas the zTh-1 – zTh-2 score was higher in the milder MDMD group than in controls. The alternative M2 profile was significantly higher in MDMD patients than in controls, while Th-17 was significantly increased in the severe MDMD group as compared with controls and the milder MDMD group.

**ESF, Figure 1** shows the mean (SE) z values of all cytokine levels (z values with the mean value of controls set at 0) in both the milder and severe MDMD groups. sIL-1RA (p<0.001), sIL-2R (p<0.001), IL-6 (p=0.002), IL-9 (p<0.001), IL-16 (p<0.001), IL-18 (p<0.001), TNF-α (p=0.001), TNF-β (p<0.001), and TRAIL (p<0.001) were significantly higher in MDMD than in controls. There was a trend towards lowered CCL3 in MDMD patients as compared with controls (p=0.306). ESF, Figure 1 shows that IL-1β (p=0.043), sIL-2R (p<0.001), IL-4 (p<0.001), IL-6 (p=0.023), IL-16 (p=0.001), IL-17 (p=0.005), IL-18 (p=0.029), MIF (p=0.004), and TRAIL (p=0.018) were significantly higher in severe MDMD than in milder MDMD.

**Figure 1.**
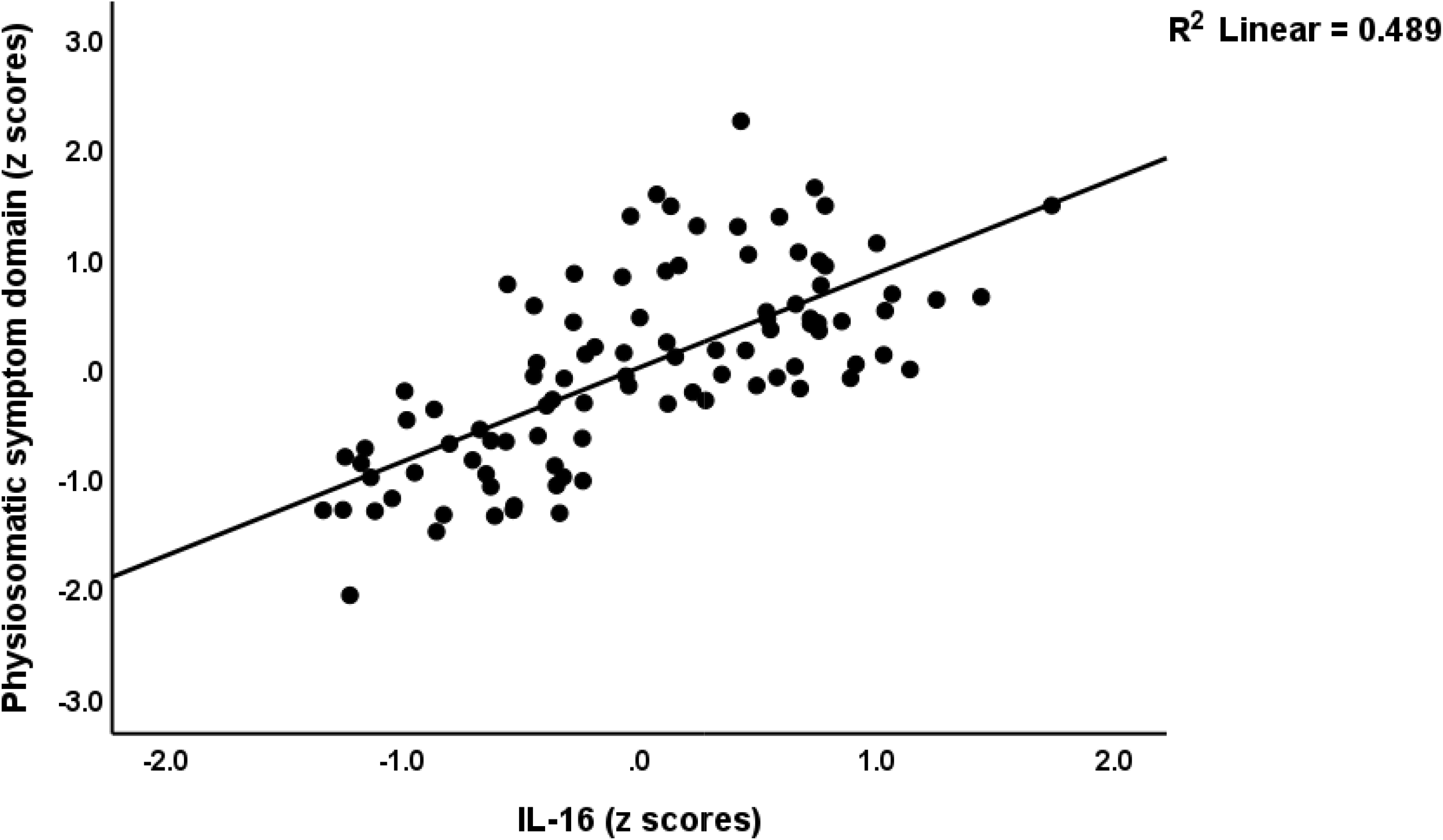
Partial regression of the physiosomatic symptom domain on interleukin (IL)-16 (p<0.001).

**ESF, Figure 2** shows the mean (SE) z values of all chemokines/growth factor levels (z values with the mean value of controls set at 0) in the milder and severe MDMD groups. The following chemokines/growth factors were significantly higher in patients than controls: CCL2 (p<0.001), CCL4 (p<0.001), CCL5 (p<0.001), CCL11 (p<0.001), CCL27 (p<0.001), CXCL1 (p<0.001), CXCL9 (p<0.001), CXCL10 (p<0.001), M-CSF (p<0.001), GM-CSF (p=0.012), SCF (p<0.001), SCGF (p<0.001), SDF1 (p<0.001), NGF (p=0.011), PDGF (p<0.001), HGF (p<0.001). We found that CCL2 (p=0.013), CCL7 (p=0.036), CCL11 (p=0.012), CCL27 (p<0.001), CXCL9 (p=0.007), M-CSF (p=0.006), SCF (p=0.001), SCGF (p=0.006), and FGF (p=0.039) were significantly higher in severe MDMD than in milder MDMD.

**Figure 2.**
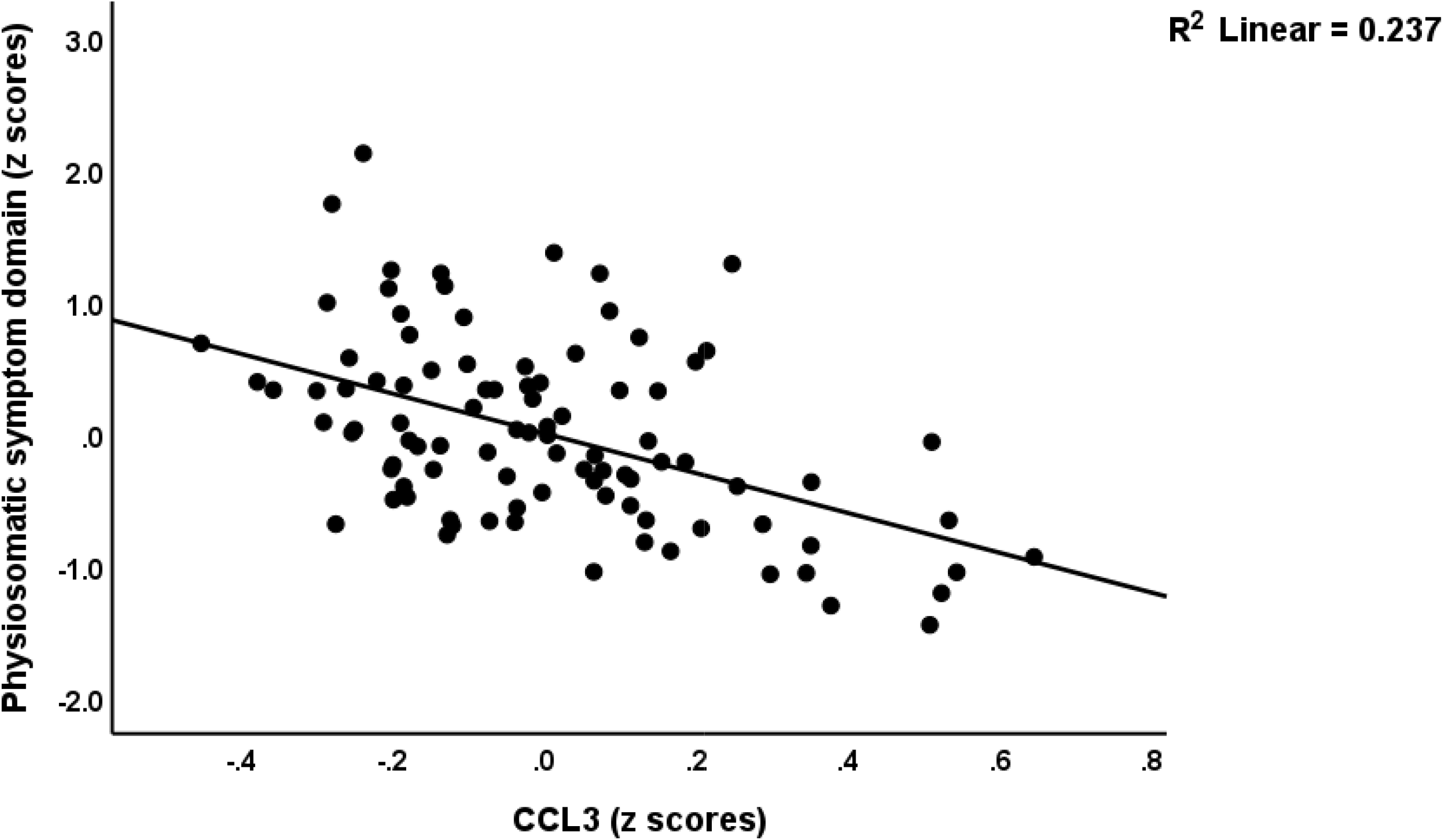
Partial regression of the physios matic symptom domain on CCL3 (p<0.001).

**Figure 3.**
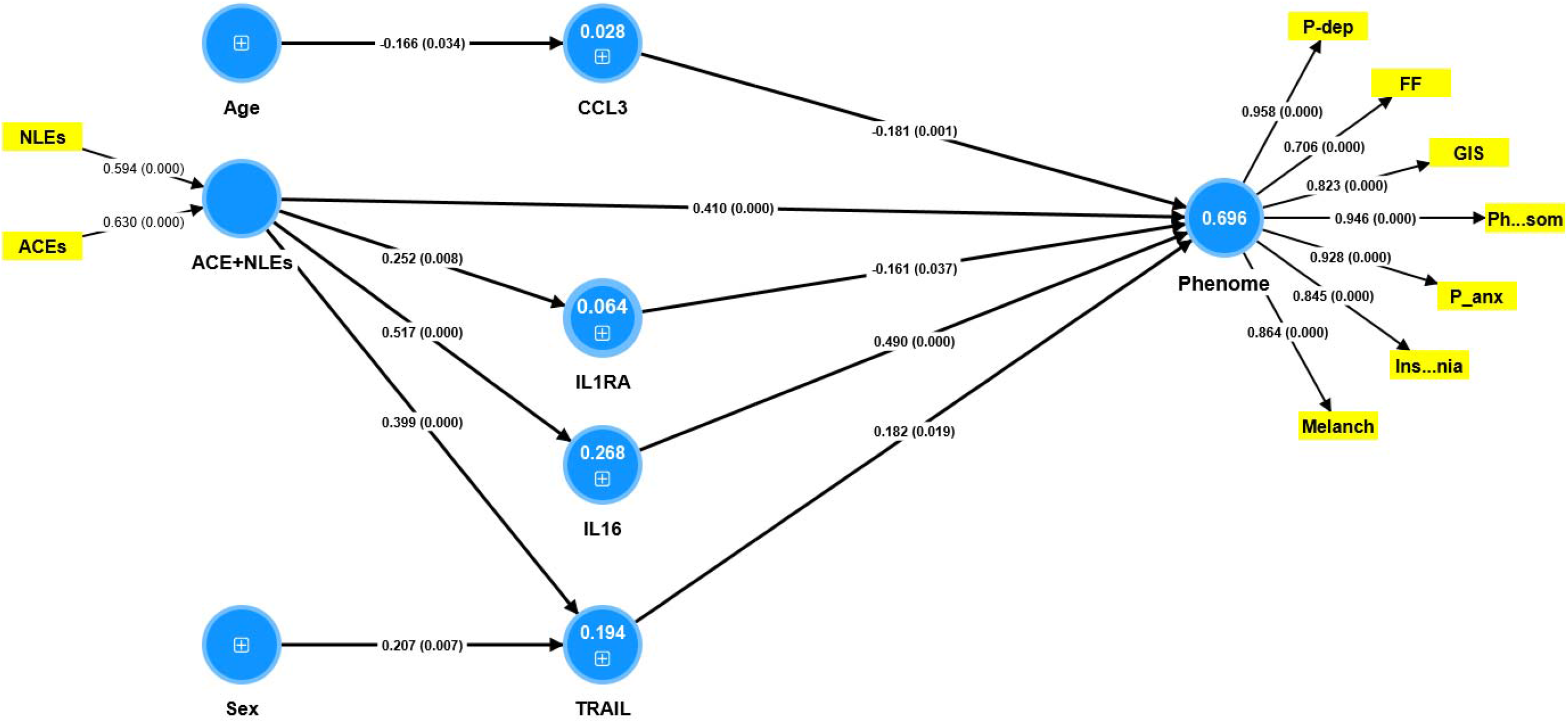
Results of Partial Least Squares analysis. The phenome of depression was conceptualized as a latent vector extracted from physiosomatic (Ph..som) and Fibro-fatigue (F symptoms, as well as pure depression (P-dep), pure anxiety (P-anx), gastro-intestinal (GIS) symptoms, melancholia (Melanch), insomnia symptoms. Adverse childhood experiences + negative life events (ACEs+NLEs), interleukin (IL)-16, TNF-related apoptosis-inducing ligand (TRAIL), soluble IL-1 receptor antagonist (sIL-1RA) and CCL3 predict 69.6% of the variance in the physio-af phenome of depression. Part of the variance in IL-16 (26.8%), TRAIL (19.4%) and sIL-1RA (6.4%) is explained by ACE+NLE Shown are path coefficients (p values), loadings (p values) or weights (p values). Figures in blue circles: variance explained.

### Intercorrelation matrix

**Table 3** shows the correlation between the major symptom domains and the immune profiles. There were no significant associations between any of the immune profiles and autonomic symptoms. The zM1 – zM2 and zIRS – zCIRS scores were not correlated with all symptom domains. The M1 classical, Th-1, IRS, CIRS, and Immu-NT profiles were significantly correlated with all symptom domains (except autonomic symptoms). The alternative M2 profile was associated with all symptom domains, except pure FF, GIS, and autonomic symptoms. Th-2 was significantly correlated with pure_depression, pure_anxiety, melancholia, and insomnia. Th-17 was associated with all symptom domains except pure_FF and autonomic symptoms. The zTh-1 – zTh-2 profile was significantly correlated with all domains, except melancholia, insomnia, and autonomic symptoms.

**Table 3.** Intercorrelation matrix between immune profiles and symptoms domain scores.

| **Variables** | **Pure depression** | **Pure anxiety** | **Pure FF** | **Physiosomatic** | **Melancholia** | **GIS** | **Insomnia** | **Autonomic** |
| --- | --- | --- | --- | --- | --- | --- | --- | --- |
| Classical M1 | 0.417 ** | 0.393 ** | 0.229 * | 0.295 * | 0.398 ** | 0.283 ** | 0.347 ** | 0.147 |
| Alternative M2 | 0.332 ** | 0.297 ** | 0.171 | 0.230 * | 0.306 ** | 0.188 | 0.269 ** | 0.112 |
| zM1 – zM2 | 0.140 | 0.157 | 0.095 | 0.157 | 0.149 | 0.155 | 0.128 | 0.057 |
| Thelper (Th)-1 | 0.405 ** | 0.365 ** | 0.252 * | 0.328 ** | 0.397 ** | 0.313 ** | 0.356 ** | 0.172 |
| Th-2 | 0.228 * | 0.215 * | 0.132 | 0.196 | 0.296 ** | 0.169 | 0.255 * | 0.187 |
| zTh-1 – zTh-2 | 0.332 ** | 0.282 ** | 0.224 * | 0.249 * | 0.189 | 0.270 ** | 0.190 | 0.029 |
| Th-17 | 0.242 * | 0.238 * | 0.135 | 0.204 * | 0.267 ** | 0.213 * | 0.242 * | 0.181 |
| IRS | 0.424 ** | 0.406 ** | 0.263 * | 0.331 ** | 0.410 ** | 0.338 ** | 0.389 ** | 0.157 |
| CIRS | 0.364 ** | 0.349 ** | 0.240 * | 0.300 ** | 0.360 ** | 0.261 * | 0.368 ** | 0.129 |
| zIRS - zCIRS | 0.142 | 0.137 | 0.053 | 0.074 | 0.120 | 0.056 | 0.049 | 0.067 |
| Immu-NT | 0.375 ** | 0.360 ** | 0.235 * | 0.300 ** | 0.367 ** | 0.312 ** | 0.368 ** | 0.151 |

\*p<0.05, \*\*p<0.001
IRS: immune-inflammatory response system, CIRS: compensatory immunoregulatory system, Immu-NT: immune-associated neurotoxicity.
FF: Fibro-Fatigue, GIS: gastro-intestinal symptoms.

### Results of multiple regression analysis

**Table 4** shows the results of different multiple regression analyses with the symptom domains as dependent variables and the cytokines/chemokines/growth factors as dependent variables, while allowing for the effects of age, sex, smoking, education, and BMI. Since we found that multiple CIRS cytokines were significant in the final regression analysis we also entered the CIRS score as an additional explanatory variable. Regression #1 shows that 48.9% of the variance in the pure_depression score was explained by IL-16 and SCGF (both positively) and CCL3 (inversely). Regression #2 shows that 45.4% of the variance in the pure_anxiety score was explained by IL-16 and IL-17 (both positively) and CCL3 (inversely). Regression #3 shows that 59.0% of the variance in physiosomatic symptoms was explained by IL-16, IL-8, and male sex (all positively associated) and CCL3 and sIL-1RA (both inversely). **Figure 1** and **Figure 2** show the partial regression of the physiosomatic symptom domain on IL-16 and CCL3, respectively. We found that 46.5% of the variance in the pure_FF score was explained by IL-16 and TRAIL (positively) and inversely by the CIRS score (regression #4). We found that 53.2% of the variance in melancholia was explained by the regression on IL-16 and SCGF (both positively) and CCL3 (inversely). A large part of the variance in GIS was predicted by IL-16, CCL1, TRAIL, and IL-8 (all positively) and CCL3 (inversely). Up to 54.1% of the variance in the insomnia score was predicted by IL-16 and SCGF (both positively) and sIL-1RA (inversely). Only a small part of the variance in autonomic symptoms was explained by TRAIL.

**Table 4.** Results of multiple regression analysis with symptom domains as dependent variables and cytokines/chemokines/growth factors as explanatory variables.

| Dependent Variables | Explanatory Variables | Coefficients of input variables |  |  | Model statistics |  |  |  |
| --- | --- | --- | --- | --- | --- | --- | --- | --- |
| | | $\beta$ | t | p | R<sup>2</sup> | F | df | p |
| #1. Pure depression | Model |  |  |  | 0.489 | 29.29 | 3/92 | <0.001 |
|  | IL-16 | 0.593 | 6.87 | <0.001 |  |  |  |  |
|  | CCL3 | -0.453 | -0.38 | <0.001 |  |  |  |  |
|  | SCGF | 0.275 | 2.16 | 0.033 |  |  |  |  |
| #2 Pure anxiety | Model |  |  |  | 0.454 | 25.55 | 3/92 | <0.001 |
|  | IL-16 | 0.593 | 6.87 | <0.001 |  |  |  |  |
|  | CCL3 | -0.453 | -3.83 | <0.001 |  |  |  |  |
|  | IL-17 | 0.275 | 2.16 | 0.033 |  |  |  |  |
| #3. Physiosomatic symptoms | Model |  |  |  | 0.590 | 25.88 | 90/5 | <0.001 |
|  | IL-16 | 0.861 | 9.28 | <0.001 |  |  |  |  |
|  | CCL3 | -0.398 | -4.93 | <0.001 |  |  |  |  |
|  | sIL-1RA | -0.266 | -2.85 | 0.005 |  |  |  |  |
|  | Sex | 0.266 | 3.58 | <0.001 |  |  |  |  |
|  | IL-8 | 0.252 | 3.09 | 0.003 |  |  |  |  |
| #4. Pure FF | Model |  |  |  | 0.465 | 26.63 | 3/92 | <0.001 |
|  | IL-16 | 0.736 | 5.68 | <0.001 |  |  |  |  |
|  | CIRS | -0.468 | -4.10 | <0.001 |  |  |  |  |
|  | TRAIL | 0.269 | 2.27 | 0.026 |  |  |  |  |
| #5. Melancholia | Model |  |  |  | 0.532 | 34.92 | 3/92 | <0.001 |
|  | IL-16 | 0.492 | 4.72 | <0.001 |  |  |  |  |
|  | SCGF | 0.306 | 3.02 | 0.003 |  |  |  |  |
|  | CCL | -0.197 | -2.63 | 0.010 |  |  |  |  |
| #6. GIS | Model |  |  |  | 0.494 | 17.55 | 5/90 | <0.001 |
|  | IL-16 | 0.305 | 2.47 | 0.016 |  |  |  |  |
|  | CCL3 | -0.395 | -4.59 | <0.001 |  |  |  |  |
|  | CCL1 | 0.310 | 3.71 | <0.001 |  |  |  |  |
|  | TRAIL | 0.271 | 2.35 | 0.021 |  |  |  |  |
|  | IL-8 | 0.171 | 2.04 | 0.044 |  |  |  |  |
| #7. Insomnia | Model |  |  |  | 0.541 | 36.17 | 3/92 | <0.001 |
|  | IL-16 | 0.676 | 5.57 | <0.001 |  |  |  |  |
|  | sIL-1RA | -0.276 | -2.85 | 0.005 |  |  |  |  |
|  | SCGF | 0.271 | 2.75 | 0.007 |  |  |  |  |
| #8. Autonomic | Model |  |  |  | 0.057 | 5.65 | 1/94 | 0.019 |
|  | TRAIL | 0.238 | 2.38 | 0.019 |  |  |  |  |

IL: interleukin, SCGF: stem cell growth factor, sIL-1RA: soluble IL-1 receptor antagonist, sex: males=1, females=0, CIRS: compensatory immunoregulatory system, TRAIL: tumor necrosis factor-related apoptosis-inducing ligand.

### Effects of ACEs and NLEs on the phenome are mediated by immune aberrations

Since there are associations between ACE+NLEs and the immune profiles as well as symptom domains (Almulla et al., 2023), we also examined whether the regression of the symptoms domains on the immune products was influenced by ACE+NLEs. **Table 5** shows the results of multiple regression analyses which included the significant effects of ACE+NLEs on the phenome data. Thus, the ACE+NLEs score had a significant effect on all symptom domains, except autonomic symptoms. The results of this Table also show that cytokines/chemokines/growth factors had significant effects on the symptom domains beyond the effects of ACE+NLEs. In all cases, the explained variances were significantly enhanced by entering the ACE+NLEs values with around 3% (variance explained), except for GIS (no significant improvement in explained variance).

**Table 5.** Results of multiple regression analysis with symptom domains as dependent variables and cytokines/chemokines/growth factors, adverse childhood experiences (ACEs) and negative life events (NLEs) as explanatory variables.

| Dependent Variables | Explanatory Variables | Coefficients of input variables |  |  | Model statistics |  |  |  |
| --- | --- | --- | --- | --- | --- | --- | --- | --- |
| | | $\beta$ | t | p | R<sup>2</sup> | F | df | p |
| #1. Pure depression | **Model** |  |  |  | 0.520 | 24.63 | 4/91 | <0.001 |
|  | IL-16 | 0.523 | 5.67 | <0.001 |  |  |  |  |
|  | CCL3 | -0.313 | -3.88 | <0.001 |  |  |  |  |
|  | ACE+NLEs | 0.219 | 2.43 | 0.017 |  |  |  |  |
|  | SCGF | 0.171 | 2.19 | 0.031 |  |  |  |  |
| #2 Pure anxiety | **Model** |  |  |  | 0.503 | 23.03 | 4/91 | <0.001 |
|  | IL-16 | 0.509 | 5.43 | <0.001 |  |  |  |  |
|  | ACE+NLEs | 0.281 | 3.10 | 0.003 |  |  |  |  |
|  | CCL3 | -0.222 | -2.81 | 0.006 |  |  |  |  |
|  | Education | -0.149 | -1.99 | 0.049 |  |  |  |  |
| #2. Physiosomatic symptoms | **Model** |  |  |  | 0.620 | 24.24 | 6/89 | <0.001 |
|  | IL-16 | 0.709 | 6.68 | <0.001 |  |  |  |  |
|  | ACE+NLEs | 0.225 | 2.68 | 0.009 |  |  |  |  |
|  | CCL3 | -0.321 | -3.87 | <0.001 |  |  |  |  |
|  | Sex | 0.255 | 3.55 | 0.001 |  |  |  |  |
|  | sIL-1RA | -0.231 | -2.53 | 0.013 |  |  |  |  |
|  | IL-8 | 0.190 | 2.32 | 0.023 |  |  |  |  |
| #4. Pure FF | **Model** |  |  |  | 0.498 | 22.58 | 4/91 | <0.001 |
|  | IL-16 | 0.715 | 5.50 | <0.001 |  |  |  |  |
|  | CIRS | -0.365 | -3.18 | 0.002 |  |  |  |  |
|  | ACE+NLEs | 0.248 | 2.70 | 0.008 |  |  |  |  |
|  | Sex | 0.164 | 2.14 | 0.035 |  |  |  |  |
| #5. Melancholia | **Model** |  |  |  | 0.568 | 40.36 | 3/92 | <0.001 |
|  | IL-16 | 0.320 | 3.27 | 0.002 |  |  |  |  |
|  | ACEQ24710 | 0.310 | 3.89 | <0.001 |  |  |  |  |
|  | SCGF | 0.247 | 2.80 | 0.006 |  |  |  |  |
| **#6. GIS** | **Model** |  |  |  | 0.494 | 17.55 | 5/90 | <0.001 |
|  | CCL3 | -0.431 | -4.60 | <0.001 |  |  |  |  |
|  | CCL11 | 0.316 | 3.83 | <0.001 |  |  |  |  |
|  | ACE+NLEs | 0.202 | 2.41 | 0.018 |  |  |  |  |
|  | M-CSF | 0.350 | 3.97 | <0.001 |  |  |  |  |
|  | IL-8 | 0.238 | 2.82 | 0.006 |  |  |  |  |
|  | CXCL1 | 0.218 | 2.46 | 0.016 |  |  |  |  |
| **#7. Insomnia** | **Model** |  |  |  | 0.570 | 30.21 | 4/91 | <0.001 |
|  | IL-16 | 0.568 | 4.52 | <0.001 |  |  |  |  |
|  | ACE+NLEs | 0.208 | 2.49 | 0.015 |  |  |  |  |
|  | sIL-1RA | -0.246 | -2.59 | 0.011 |  |  |  |  |
|  | SCGF | 0.234 | 2.41 | 0.018 |  |  |  |  |

IL: interleukin, SCGF: stem cell growth factor, sIL-1RA: soluble IL-1 receptor antagonist, sex: males=1, females=0, CIRS: compensatory immunoregulatory system, TRAIL: tumor necrosis factor-related apoptosis-inducing ligand.

### Results of PLS analysis

The first PLS model conceptualized the phenome of depression as a latent vector extracted from physiosomatic and FF symptoms, as well as pure_depression, pure_anxiety, GIS symptoms, melancholia, and insomnia. The ACEs+NLEs index was composed as a formative model. Based on the results of the multiple regression and correlation analyses, we entered the most significant cytokines/chemokines/growth factors predicting the phenome, and in addition included the effects of ACE+NLEs on the immune variables (together with age and sex). The composite reliability (0.956), Cronbach’s alpha (0.945) and AVE (0.759) of the phenome latent vector were more than adequate, as was the model quality fit with an SRMR of 0.047. All Q^2^ predicted values for the manifest and latent variables were positive using PLSpredict, indicating that the model outperforms the naivest benchmark. We observed that the phenome of depression was best predicted by ACE+NLEs, IL-16, TRAIL (all positively), and sIL-1RA and CCL3 (both inversely). The latter five predictors explained 69.6% of the variance in the phenome of depression. In addition, part of the variance in IL-16 (26.8%), TRAIL (19.4%) and sIL-1RA (6.4%) was explained by ACE+NLEs. Age was associated with CCL3 and sex with TRAIL. As such, there were significant specific indirect effects of ACE+NLEs on the phenome which were partly mediated by IL-16 (t=4.40, p<0.001) and TRAIL (t=2.16, p=0.040). The direct and total indirect (t=5.99, p<0.001) effects of ACE+NLEs on the phenome explain the strong total effects of ACE+NLEs on the phenome (t=13.21, p<0.001).

Consequently, we have constructed a second PLS model by integrating different cytokines/chemokine/growth factors into one latent vector, labeled “immune network,” including IL-16, TRAIL, CCL27, M-CSF, and SCGF. This factor showed adequate convergence and construct validity as indicated by an AVE value of 0.736, composite reliability of 0.933, and Cronbach’s alpha of 0.910. Moreover, the overall model fit was adequate with an SRMR value of 0.043. We found that 68.8% of the variance in the phenome of depression was explained by this immune network (positively associated) and CCL3 (inversely associated). ACE+NLEs had significant effects on the phenome, comprising direct effects (see Figure 4) as well as specific indirect effects (t=6.55, p<0.001) which were partially mediated by the immune network. Both effects of ACE+NLEs culminated in strong total effects (t=13.41, p<0.001). On the other hand, there was no significant effect of age on the phenome (t=1.62, p=0.106).

**Figure 4.**
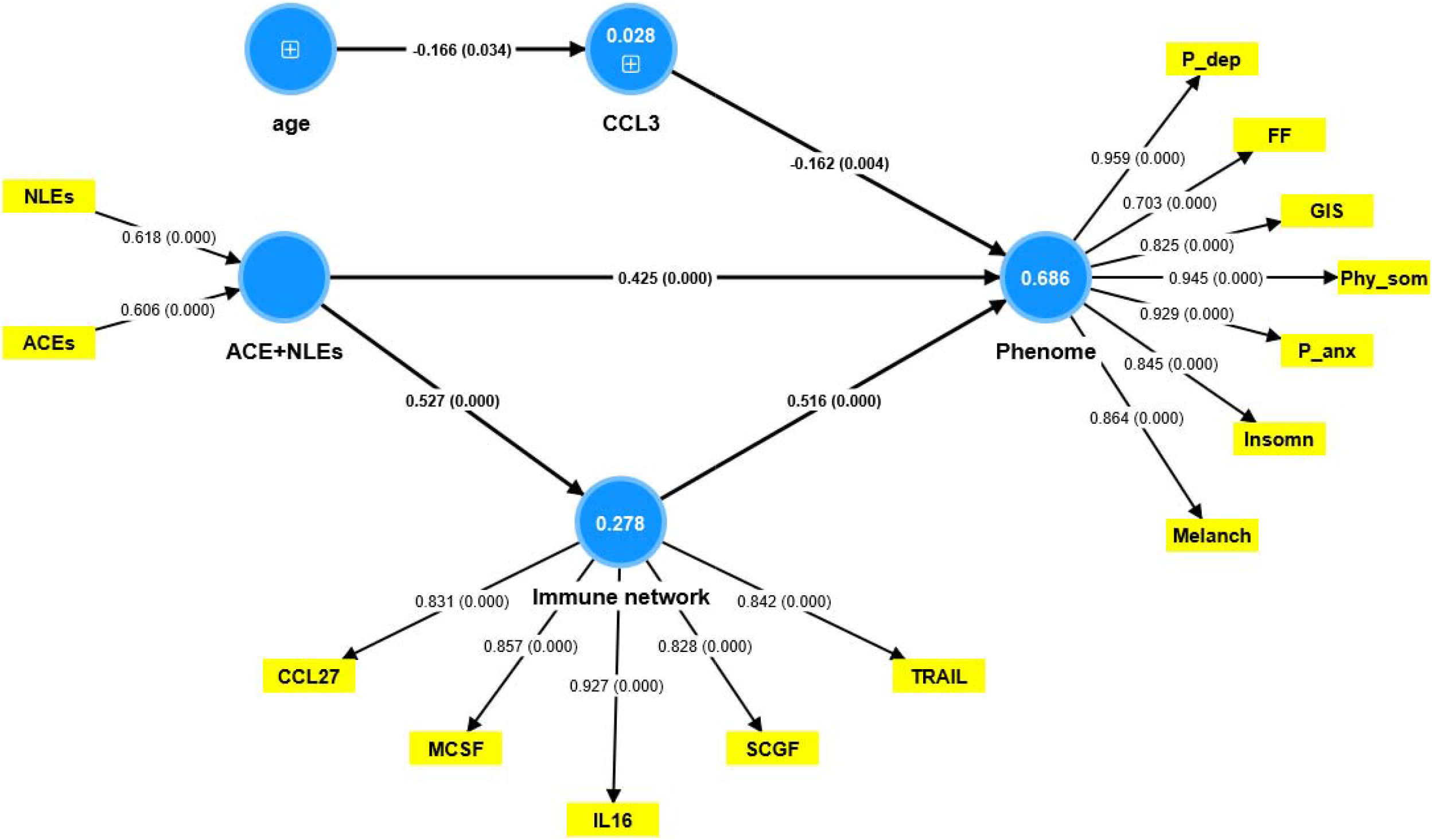
Results of Partial Least Squares analysis. The phenome of depression was conceptualized as a latent vector extracted from physiosomatic (Ph..som) and Fibro-fatigue (FF) symptoms, as well as pure depression (P-dep), pure anxiety (P-anx), gastro-intestinal (GIS) symptoms, melancholia (Melanch), and insomnia. Adverse childhood experiences + negative life events (ACEs+NLEs), CCL3, and a factor extracted from 5 cytokines/chemokines/growth factors (labeled: immune network) explained 68.6% of the variance in the physio-affective phenome. IL-16: interleukin (IL)-16, TRAIL: TNF-related apoptosis-inducing ligand, M-CSF: macrophage colony stimulating factor, SCGF: stem cell growth factor. Shown are path coefficients (p values) and loadings (p values) or weights (p values). Figures in blue circles: variance explained.

## Discussion

### The physio-affective phenome of depression

The first major discovery of this study indicates that within MDMD patients, the physiosomatic, FF, and GIS symptom domains are associated with the same factor as pure depression, pure anxiety, melancholia symptoms, and insomnia. This suggests that the various symptom domains are closely interconnected expressions of a same latent construct, specifically the phenome of depression. Previous studies have demonstrated that physiosomatic symptoms are associated with the same phenome factor as pure depressive and anxiety symptoms (Anderson, Berk et al. 2014, Al-Hakeim, Al-Naqeeb et al. 2023). To emphasize the influence of interconnected affective and physiosomatic symptoms on the manifestation of MDD, this construct has been renamed as the physio-affective phenome (Maes, Al-Rubaye et al. 2022, Al-Hakeim, Al-Naqeeb et al. 2023). Furthermore, various medical and neuro-psychiatric disorders exhibit a latent construct encompassing symptoms of depression, anxiety, and physiosomatic manifestations. This phenomenon can be observed in conditions such as acute COVID-19 infection (Al-Jassas, Al-Hakeim et al. 2022), Long COVID (Maes, Al-Rubaye et al. 2022), relapsing-remitting multiple sclerosis (RRMS) (Almulla, Abdul Jaleel et al. 2023), schizophrenia (Kanchanatawan, Thika et al. 2018), and epilepsy (Maes, Supasitthumrong et al. 2020). Furthermore, FF symptoms belong to the same latent construct which loads highly on affective and physiosomatic symptoms. The aforementioned interconnected relationships among FF symptoms and the phenome of depression were previously identified in the context of MDD (Al-Hakeim, Al-Naqeeb et al. 2023), acute COVID-19 infection (Almulla, Al-Hakeim et al. 2023), RRMS (Almulla, Abdul Jaleel et al. 2023), Long COVID (Al-Hakeim, Al-Rubaye et al. 2022), schizophrenia (Maes, Andrés-Rodríguez et al. 2023), and end-stage renal disease (Asad, Al-Hakeim et al. 2023).

The interrelationship between affective, physiosomatic, and FF symptoms is well-established, indicating that these symptoms are interconnected and should not be considered distinct phenomena. This implies that MDD, bipolar disorder, and various psychiatric disorders exhibit a physio-affective phenome. These findings also suggest that the physiosomatic, FF and GIS symptom domains, as well as pure depression and anxiety, melancholia, and insomnia, are associated with similar underlying mechanisms that involve both central and peripheral aberrations. This observation suggests that MDMD is a condition that affects the entire body and that affective symptoms are merely one component of the comprehensive clinical presentation.

### The physio-affective phenome is predicted by immune activation

The results of the present study show that physiosomatic and FF symptoms of MDMD are best predicted by a) IRS, Th-1 activation, increased Immu-NT, Th-1 polarization, and M1 activation; b) a combination of increased IRS products (especially IL-16, IL-8 or TRAIL), lowered CIRS components (sIL-1RA), and lowered levels of a chemokine (CCL3), and c) an immune network comprising IL-16, TRAIL, CCL27, M-CSF and SCGF (all positively) and CCL3 (inversely). These results indicate that T cell activation (IL-16, Th-1 polarization) is a significant factor in the development of physiosomatic, FF, and GIS symptoms.

IL-16 is elevated in major depressive disorder (Timothy, Helena et al. 2018) and is a crucial cytokine in FE-MDMD (Almulla, Ali Abbas Abo et al. 2023a). IL-16 and other proinflammatory cytokines are associated with elevated depression and neuroticism scores in hepatitis C patients receiving cytokine-based immunotherapy (Mathy, Scheuer et al. 2000, Pawlowski, Radkowski et al. 2014, Hall, Cullen et al. 2016). IL-16 induces T cell signaling via the CD4 molecule on T helper cells, resulting in the upregulation of activation markers such as HLA-DR+ and CD25+ cells. Prenatal depression is associated with elevated IL-16 levels in the prefrontal cortex and hippocampus in animal models (Posillico and Schwarz 2016) and contributes to neuroinflammation in animal models of autoimmune encephalomyelitis by activating T cells (Hridi, Barbour et al. 2021). CCL27 or CTACK (cutaneous T cell-attracting chemokine) is a pro-inflammatory chemokine that attracts memory T cells (T lymphocyte-associated antigen) and this chemokine is essential for T cell-mediated cutaneous inflammation (Morales, Homey et al. 1999, Homey, Alenius et al. 2002). Such interactions may explain the co-occurrence of depression and “psychocutaneous disorders” like psoriasis, which is accompanied by elevated CCL27 levels (Ye, Ren et al. 2017). Moreover, CCL27 is a key regulator of immune homeostasis in mucosal tissues and the epidermis (Davila, Xu et al. 2022), and CCL27 KO-mice exhibit CR10+ T cell infiltration in the reproductive tract and lungs (Davila, Xu et al. 2022).

Another cytokine that is strongly associated with physiosomatic and FF symptoms is TRAIL. TRAIL signals apoptosis by binding to death receptors and activating the caspase pathway (Chyuan and Hsu 2020). This cytokine has strong neurotoxic effects, even leading to neurodegeneration and neuronal death, mediated by the TRAIL-R2/DR5 receptor and caspase activation (Di Benedetto, Burgaletto et al. 2022). Recent evidence suggests, however, that TRAIL may modulate the IRS and T cell activation and shows anti-inflammatory effects (Chyuan and Hsu 2020). TRAIL inhibits the activation of T cells, indicating that this cytokine functions as an activator of an anti-inflammatory pathway independent of apoptosis.

Many patients with MDD exhibit T cell activation, as measured by flow cytometry and T cell activation markers including HLA-DR+ and CD7+CD25+ T cells (Maes, Lambrechts et al. 1992, Maes, Stevens et al. 1993). We have argued, based on these and other findings, that cell-mediated activation, and specifically T cell activation, is a key factor in MDD (Maes 2011). CD3+CD71+, CD3+HLADR+, and CD4+CD71+ T cell phenotypes have been demonstrated to differentiate MDMD from SDMD (Maes, Rachayon et al. 2023). In addition, the numbers of CD3+CD40L, CD4+CD40L, CD4+HLADR+, and CD8+CD40L+ cells are substantially higher in MDMD than in healthy controls, whereas SDMD patients occupy an intermediate position (Maes, Rachayon et al. 2023).

Importantly, physiosomatic and FF symptoms are strongly associated with Th-1 cytokine profiles, but not with Th-2 profiles. Th-2 cytokines (such as IL-4) have negative immune-regulatory and anti-inflammatory effects and are essential for CIRS activity. As a result, a relative decrease in Th-2 functions may result in attenuated immunoregulatory Th-2 effects, thereby enhancing the deleterious effects of Th-1 and IRS activation. Similarly, our results demonstrate that decreased CIRS activity plays a crucial role in FF symptoms. Even more significant is the fact that relative decreases in sIL-1RA levels (another component of CIRS) contribute to physiosomatic symptoms and the physio-affective phenome of depression. There is evidence that both MDD and MDMD are associated with elevated sIL-1RA serum/plasma concentrations (Maes, Smith et al. 1995, Almulla, Ali Abbas Abo et al. 2023a). Maes and Carvalho (2018) report that soluble sIL-RA may inhibit both IL-1α and IL-1β signaling, thereby exerting potent anti-inflammatory effects. Recent findings indicate that the phenome of MDMD is significantly predicted by activated T and B cells, whereas T regulatory cells (such as CD25+FoxP3+GARP+) are inversely associated with the phenome (Maes, Rachayon et al. 2023). Based on the IRS/CIRS theory of depression (Maes and Carvalho 2018), we may conclude that increased Th-1 (IL-16), TRAIL and M1 activities, and relative deficiencies in CIRS (including Th-2 cytokines and sIL-1RA) are major contributors to physiosomatic and FF symptoms.

The analyses that we conducted in the current study suggest that a decrease in CCL3 may contribute to the severity of physiosomatic and FF symptoms, which is another potentially significant finding. CCL3 (or macrophage inflammatory protein) is a pro-inflammatory chemokine that recruits activated immune cells to inflammatory sites and increases the expression of pro-inflammatory M1 cytokines including IL-1β, TNF-α, and IL-6 (Zhang, Qiao et al. 2018). Previously, we found that whole blood-stimulated CCL3 production in the culture supernatant of patients with MDMD was significantly lower than that of healthy controls (Rachayon, Jirakran et al. 2023). In addition, CCL3 was one of the few cytokines/chemokines whose serum concentrations were lower in the serum of MDMD patients compared with controls (Almulla, Ali Abbas Abo et al. 2023a). Consequently, we can hypothesize that relative decreases in CCL3 secretion may play a role in the depression phenome. CCL3 mediates fever independent of prostaglandin (PGE2) and elevates PGE2 in cerebrospinal fluid (Miñano, Sancibrian et al. 1990, Soares, Hiratsuka Veiga-Souza et al. 2006, Soares, Figueiredo et al. 2009). Endogenous PGE2 possesses context-dependent anti-inflammatory properties and contributes to the resolution of inflammation (Scher and Pillinger 2009, Frolov, Yang et al. 2013). In addition, CCL3 has neuroprotective properties, such as protecting dopaminergic neurons, and influencing traumatic brain injuries (Leggio, L’Episcopo et al. 2022).

### Effects of ACE and NLEs on the phenome are mediated by the immune network

The third major finding is that the combined independent effects of immune aberrations and ACE+NLEs significantly predict the physiosomatic and FF symptoms of MDMD, and the effects of ACE+NLEs on the phenome are partially mediated by increased IL-16 and TRAIL, whereas ACE-activation of sIL-1RA levels may have protective effects. Our second PLS analysis revealed that ACE+NLEs exert a substantial influence on an immune network composed of IL-16, CCL27, TRAIL, MCSF, and SCGF, which partially mediates the effects of ACE+NLEs on the phenome, including physiosomatic and FF symptoms. SCGF (CLECC11a) activates different hematopoietic progenitor cells and activates cell signaling networks including glycogen synthase kinase 3, β-catenin and Wnt pathways, which play a major role in MDD, physiosomatic and FF symptoms (Maes, Fišar et al. 2012, Maes, Rachayon et al. 2022). M-CSF is a macrophage differentiation factor that is essential for microglial activation and neuroinflammation (Ghia, Galeazzi et al. 2008, Smith, Gibbons et al. 2013).

Together, ACEs and NLEs appear to induce a pro-inflammatory network composed of cytokines, chemokines, and growth factors. These findings extend those of a previous paper (Maes, Rachayon et al. 2022), which demonstrated that ACEs elicit a network of cytokines/chemokines/growth factors measured in the culture supernatant of stimulated whole blood from MDMD patients. In fact, both the present investigation (conducted on Iraqi patients) and our previous report (conducted on Thai patients) (Maes, Rachayon et al. 2022) demonstrate that ACEs activate M1, Th-1, Th-17, IRS, and CIRS profiles and significantly increase Immu-NT. In addition, ACEs were associated with elevated levels of sIL-1RA, IL-9, IL-12p70, PDGF, and TNF-α in both studies.

Maes et al. (2022) demonstrated that ACEs induce stimulated but not unstimulated levels of cytokines/chemokines/growth factors, suggesting that the latter factors are sensitized in MDMD patients. Future stressors could then reactivate these sensitized networks, according to the hypothesis. Since NLEs increase the serum levels of IL-16, CXCL12, M-CSF, SCGF, TRAIL, IL-18, PDGF, sIL-2R, sIL-1RA, SCF, and IL-9, and ACE+NLEs have a greater effect on CXCL12 and M-CSF than ACEs, we can conclude that NLEs may indeed reactivate the sensitized ACE-induced immune responses. However, ACE+NLEs also had significant direct effects on the MDMD phenome, indicating the involvement of other pathways, such as gut-brain pathways, gut-dysbiosis, oxidative stress pathways, and decreased antioxidants and neurotrophic protection (Maes, Congio et al. 2018, Maes, Moraes et al. 2019, Maes, Rachayon et al. 2023, Maes, Vasupanrajit et al. 2023).

## Conclusions

The findings of this study indicate that physiosomatic, FF and GIS symptoms are strongly associated with the depression, anxiety, melancholia, and insomnia domains of MDMD. The first factor derived from these six different domains is designated as the physio-affective phenome of depression. A larger part of the variances in the physio-affective phenome, and the physiosomatic FF and GIS symptom domains is explained by immune variables. Specifically, IL-16, TRAIL and IL-8 have a positive correlation with these symptoms, while CCL3 and sIL-1RA exhibit an inverse correlation. The application of PLS analysis reveals that the combination of ACEs and NLEs has a significant impact on the physio-affective phenome. This influence is partially mediated by an immune network consisting of interleukin-16, CCL27, TRAIL, M-CSF, and SCGF. Overall, the physiosomatic and FF symptoms of FE-MDMD are attributed, in part, to Immu-NT linked to Th-1 polarization and M1 macrophage activation, as well as a decrease in CIRS protection. IL-16, Th-1 polarization, M1 activation and Immu-NT are new drug targets to treat the physiosomatic and FF symptoms of MDMD, and to desensitize the upregulated cytokine network and deprogram the detrimental effects of ACEs and NLEs.

## Acknowledgments

The authors acknowledge the Al-Hakeem General Hospital workers for their efforts in data collection.

## Ethical approval and consent to participate

The College of Medical Technology at The Islamic University of Najaf, Iraq (18/2021) approved the research project. Our IRB follows the International Guideline for Human Research Safety, as well as the World Medical Association Declaration of Helsinki, The Belmont Report, the CIOMS Guideline, and the International Conference on Harmonization of Good Clinical Practice, and our study was conducted in accordance with all applicable Iraqi and international ethics and privacy laws. (ICH-GCP).

## Informed Consent Statement

All participants and their parents or legal guardians signed a written consent form.

## Declaration of interest

The authors declare no conflicts of interest.

## Funding

The C2F program at Chulalongkorn University in Thailand, grant number 64.310/436/2565 to AFA, the Thailand Science Research, and Innovation Fund at Chulalongkorn University (HEA663000016), and a Sompoch Endowment Fund (Faculty of Medicine) MDCU (RA66/016) to MM all provided funding for the project.

## Author’s contributions

AFA and PS quantified the biomarkers in the blood serum. MM conducted the study’s statistical analysis. The work is written and edited by MM, AA, BZ, AAA and PS. All authors have read and approved the final manuscript.

## Availability of data

The corresponding author (MM) will make the SPSS file used in the current study available upon receipt of an appropriate request and once the author has fully exploited the data.

## Notes

### Competing Interest Statement

The authors have declared no competing interest.

### Author Declarations

The College of Medical Technology at The Islamic University of Najaf, Iraq (18/2021) approved the research project. Our IRB follows the International Guideline for Human Research Safety, as well as the World Medical Association Declaration of Helsinki, The Belmont Report, the CIOMS Guideline, and the International Conference on Harmonization of Good Clinical Practice, and our study was conducted in accordance with all applicable Iraqi and international ethics and privacy laws. (ICH-GCP). Before participating in the study, all participants, or, if applicable, their parents or legal custodians, provided informed consent in writing. Document No. 18/2021 indicates that the Ethics Committee of the College of Medical Technology at the Islamic University of Najaf in Iraq has approved this investigation.

